# Feasibility and utility of rapid antigen testing for COVID-19 in a university residence: a cross sectional study

**DOI:** 10.1101/2021.05.24.21257732

**Authors:** Sabrina T. Wong, Marc Romney, Nancy Matic, Kristen Haase, Manon Ranger, Ranjit Dhari, Frances Affleck, Elsie Tan, Innocent Ndateba, Erica Tobias, Elizabeth Saewyc, Michael Schwandt, Don Sin

**Affiliations:** University of British Columbia School of Nursing and Centre for Health Services and Policy Research, 201-2206 East Mall, Vancouver, BC Canada V6T 1Z3; Medical Microbiology and Virology, St. Paul’s Hospital/Providence Health Care and University of British Columbia Faculty of Medicine, 1081 Burrard Street Vancouver, BC Canada V6Z 1Y6; Medical Microbiology and Virology, St. Paul’s Hospital/Providence Health Care, 1081 Burrard Street, Vancouver, BC Canada V6Z 1Y6; University of British Columbia School of Nursing, 2211 West Mall, Vancouver, BC Canada V6T 2B5; Vancouver Coastal Health and University of British Columbia School of Public and Population Health, Suite 800 - 601 West Broadway, Vancouver BC V5Z 4C2; Centre for Heart Lung Innovation (HLI) and University of British Columbia Faculty of Medicine St. Paul’s Hospital/Providence Health Care, 1081 Burrard Street, Vancouver, BC

## Abstract

**Importance:** Understanding feasibility of rapid testing in congregate living setting provides critical data to reduce the risk of outbreaks in these settings.

**Objective:** Use rapid antigen screening to detect SARS-CoV-2 in an asymptomatic group of university students and staff.

**Design:** Cross-sectional

**Setting:** University of British Columbia, Vancouver, Canada.

**Participants:** Students and staff living or working in congregate housing.

**Intervention:** Health care professional administered rapid antigen test

**Main Outcomes and measures:** Use of BD Veritor rapid antigen testing and asymptomatic participants’ experiences with rapid testing

**Results:** A total of 3536 BD Veritor tests were completed in 1141 unique individuals. One third of participants completed between two to four tests and 21% were screened five or more times. The mean number of tests completed per person was three. The mean length of time between those who had more than one test was seven days. There were eight false positives and 25 PCR confirmed COVID-19 positive individuals identified through this work. All individuals reported having no symptoms that they attributed to COVID-19. Almost all (n=22, 88%) COVID-19 positive cases were found in male participants. A total of 86 additional students from multiple different student residences (n=9) were asked to self-isolate while they waited for their COVID-19 diagnostic test results. An average of seven additional students positive for COVID-19 living in congregate housing were identified through contact tracing by finding one positive case.

**Conclusions and relevance:** Rapid testing is a relatively inexpensive and operationally easy method of identifying asymptomatic individuals with COVID-19.

## Background

Over a year into the COVID-19 pandemic, testing remains a cornerstone of mitigation and containment strategies,(1) with recent data showing that one third of people infected with COVID-19 may be asymptomatic.(2) The gold standard for COVID-19 testing is reverse-transcriptase quantitative polymerase chain reaction (RT-qPCR)(3) for the detection of SARS-CoV-2. Rapid antigen tests, however, provide increased access to results at the point of care (PoC) at a lower cost and greatly improving accessibility and turnaround time making them appealing to industry, policy makers, and healthcare professionals.(3) The performance of rapid antigen tests will vary depending on population characteristics. It is important to understand the feasibility and utility of this technology in a range of settings, particularly when screening asymptomatic individuals where the risk of transmission to others is high.(4,5)

For people living in congregate housing, COVID-19 mitigation strategies are critical public health tools. In Canada, there is currently no universal (federal or provincial) public health policy for systematic COVID-19 screening for those living in congregate housing. Individuals are expected to monitor themselves daily for symptoms and use “multiple layers of protection” which include physical distancing, mask wearing, and hand washing(6). This guidance is problematic because these measures are relatively ineffective and unlikely to identify all individuals with COVID-19, especially amongst those who are minimally symptomatic or believe they are asymptomatic.(7)

Congregate living settings present a challenge for public health experts and policy makers. Though initially deemed a low risk and low prevalence setting, there is also potential for large clusters or outbreaks should transmission of the virus go undetected. Such was the case in Canadian outbreaks in two university congregate housing residence settings,(8,9) demonstrating that access to common spaces amongst large groups of people with a propensity for high social interactions has higher risk than postulated. Asymptomatic rapid testing programs have been implemented in other university settings with mixed evidence,(10–12) but successful implementation in these congregate living settings creates the opportunity to identify emerging clusters and break chains of infection earlier.(13,14)

The purpose of this pragmatic feasibility study was to determine the use of a COVID-19 rapid antigen test among people living and working in university congregate housing. The specific objectives were to:

1. Determine whether asymptomatic students and staff would volunteer to undergo semi-weekly screening; and
2. Understand the experiences of those receiving rapid antigen testing.

## Methods

We used a cross-sectional study design. We describe our work using the Wheel of intervention,(15) which has been used by public health practice as a basis for practice, teaching, and management. The “Intervention Wheel” consists of three components: 1) population-based; 2) three levels of practice (individual, community and system); and 3) identifies and defines 17 public health interventions which contribute to improving population health. The interventions are grouped into five wedges: (1) surveillance, disease and other health event investigation, outreach, screening and case finding; (2) referral and follow-up, case management and delegated functions; (3) health teaching, counseling and consultation; (4) collaboration, coalition building and community organizing; and (5) advocacy, social marketing and policy development.

Use of rapid antigen tests to detect SARS-CoV-2 (COVID-19) is considered a screening intervention. Screening tests are intended to identify infected individuals without, or prior to development of, symptoms who may be contagious so that measures can be taken to prevent further transmission. Screening is intended to identify occurrence of the condition at the individual level even if there is no reason to suspect infection. This includes, but is not limited to, screening of asymptomatic individuals without known exposure with the intent of making decisions based on the test results. Those that have tested positive on the rapid test must be assumed to have COVID-19 until proven otherwise by a confirmatory RT-PCR test. This approach of using “point-of-care” screening tests was previously advocated by Mina et. al.(16)

### Setting

This study took place at the University of British Columbia Vancouver campus in the Orchard Commons Residence. This site was chosen as a pilot site to test the feasibility of the BD Veritor COVID-19 rapid antigen test among those living or working in congregate housing. The University of British Columbia Vancouver campus had over 1500 undergraduate students living and studying in a congregate housing site during February-April, 2021.

This pilot was supported by UBC through the School of Nursing; Student Housing and Community Services which included food services, front desk, child care services, residence life; other student services including the Sexual Violence Prevention and Response Office and Student Health; Safety and Risk Services. The Vancouver Coastal Health (VCH) Medical Health Officer and VCH public health team provided support for contact tracing, case management and cluster investigation.

### Sample

Initially, we targeted a convenience sample of first-year university students and staff living or working in university housing. As positive cases were detected, we widened eligibility of the screening site to include UBC students living in specific university owned and operated housing sites (n=9), students living on campus but in privately owned housing (e.g. renting an apartment with friends, fraternity, sorority), and varsity athletes. Staff working in any of these residences or with students on an athletic team were eligible to come to the site for rapid antigen testing. People who had previously tested positive by RT-PCR for SARS-CoV-2 within the past 90 days were excluded.

### Data sources

#### Rapid antigen test

The Orchard Commons screening site employed the BD Veritor COVID-19 rapid antigen test.(17) The BD Veritor test is a lateral flow assay resembling a pregnancy test with result interpretation performed by reading by a small handheld instrument. The sample type is a bilateral nares swab. BD Veritor is known to have reduced analytical sensitivity compared to gold standard nucleic acid tests (NAT).(18) The sensitivity, as determined by prior studies ranges from 92% when the cycle threshold (Ct) cut-off was ≤ 21 for all NAT positive samples, to 61.5% when the Ct cut-off was <40. The specificity was found to be 99.5%.(19) As part of this work, we evaluated the sensitivity and specificity of the BD Veritor against the gold standard NAT, which is reported elsewhere (under review). BD Veritor tests and readers were provided by the Health Canada Testing Secretariat via a federal allocation process to Canadian provinces. Confirmatory testing by RT-PCR for RAT positives was performed on nasopharyngeal swabs by an accredited clinical virology laboratory.

#### Survey

We created a survey for participants to complete related to their experiences at the screening site and their behaviours after being identified as having COVID-19. Example questions included acceptability of the screening site, whether they would return for testing, and reasons why they got tested. The questionnaire also collected information on demographic and health characteristics of the study participants.

#### Procedures

The screening site opened on February 9, 2021, Tuesdays-Fridays. The UBC Student Housing and Community Services (SHCS) created a dedicated website describing the screening site and sent email invitations to eligible students and staff to book appointments for rapid testing. SHCS devised a communications plan to send regular emails between February and April, 2021 once the screening site was open. UBC School of Nursing faculty, staff and students administered the nasal swabs and conducted the rapid testing. All received training related to nasal swabbing. Quality control was maintained through all staff completing a written test on the BD Veritor system, quality control of the reader once per week and having a core group of staff and faculty who conducted the rapid testing in small batches. All participants were provided with their results (negative, positive, invalid). Participants were provided related health teaching as needed and encouraged to test regularly (up to two times per week). If the result was invalid, participants could either have the screening test again or come back at a time convenient for them.

If the test was positive, the registered nurse took a nasopharyngeal specimen which was couriered to the accredited laboratory for a confirmatory PCR test. Participants were asked to self-isolate until the results of the PCR test were available (within eight to ten hours). The Vancouver Coastal Health medical health officer and public health team were made aware of any confirmed positives so they could carry out contact tracing. One of the authors (SW) contacted all students with their PCR results to ensure follow-up of self-isolation and provide a source of support if it was needed.

Procedures for survey data collection were approved by the UBC Clinical Research Ethics Board [H21-00618]. Once REB approval was obtained for survey data collection, a link to the informed consent form and survey were emailed to participants when they registered for their appointment. The Qualtrics UBC survey was completed voluntarily by participants during the last three weeks of the screening site operations while they were waiting for their test results.

#### Analysis

The test and survey data were two separate de-identified datasets. We used descriptive statistics to summarize results for the test and survey data. For open ended survey questions, we categorized answers using a descriptive thematic analysis approach.(20)

## Results

The Orchard Commons screening operated between February and April 2021. During that time, 3536 BD Veritor tests were completed for a total of 1141 unique individuals. The majority of individuals visiting the screening site completed one rapid antigen test (see Table 1). One third of participants completed between two to four tests and 21% were screened five or more times. The mean number of tests completed per person was three.

**Table 1.** Number of rapid antigen tests completed per person at Orchard Commons.

| **BD Veritor rapid antigen tests** | **Participants**<br>**n (%)** |
| --- | --- |
| 1 | 514 (45) |
| 2 | 195 (17) |
| 3-4 | 188 (16) |
| 5-9 | 175 (15) |
| ≥10 | 69 (6) |

The mean length of time between those who had more than one test was seven days though this varied with almost 40% of tests occurring between 3-6 days and 32% occurring between 7-13 days (see Figure 1). Sixteen percent of tests occurred on consecutive days whereas 14% of tests were over two weeks apart.

There were 8 false positives and 25 RT-PCR confirmed SARS-CoV-2 true positives identified through the Orchard Commons screening site. For the participants who were identified as having COVID-19, all reported having no symptoms that they attributed to their positive status. Almost all (n=22, 88%) COVID-19 cases were found in male participants. Students who lived in housing owned and operated by UBC moved into self-isolation units provided by the university, and additional students were asked to self-isolate. Table 2 shows the number of additional students asked to self-isolate within three days of an Orchard Commons confirmed positive case. A total of 86 additional students from multiple different student residences (n=9) were asked to self-isolate while they waited for their COVID-19 diagnostic test results. An average of seven additional students living in congregate housing were affected by finding one positive case.

**Table 2.**
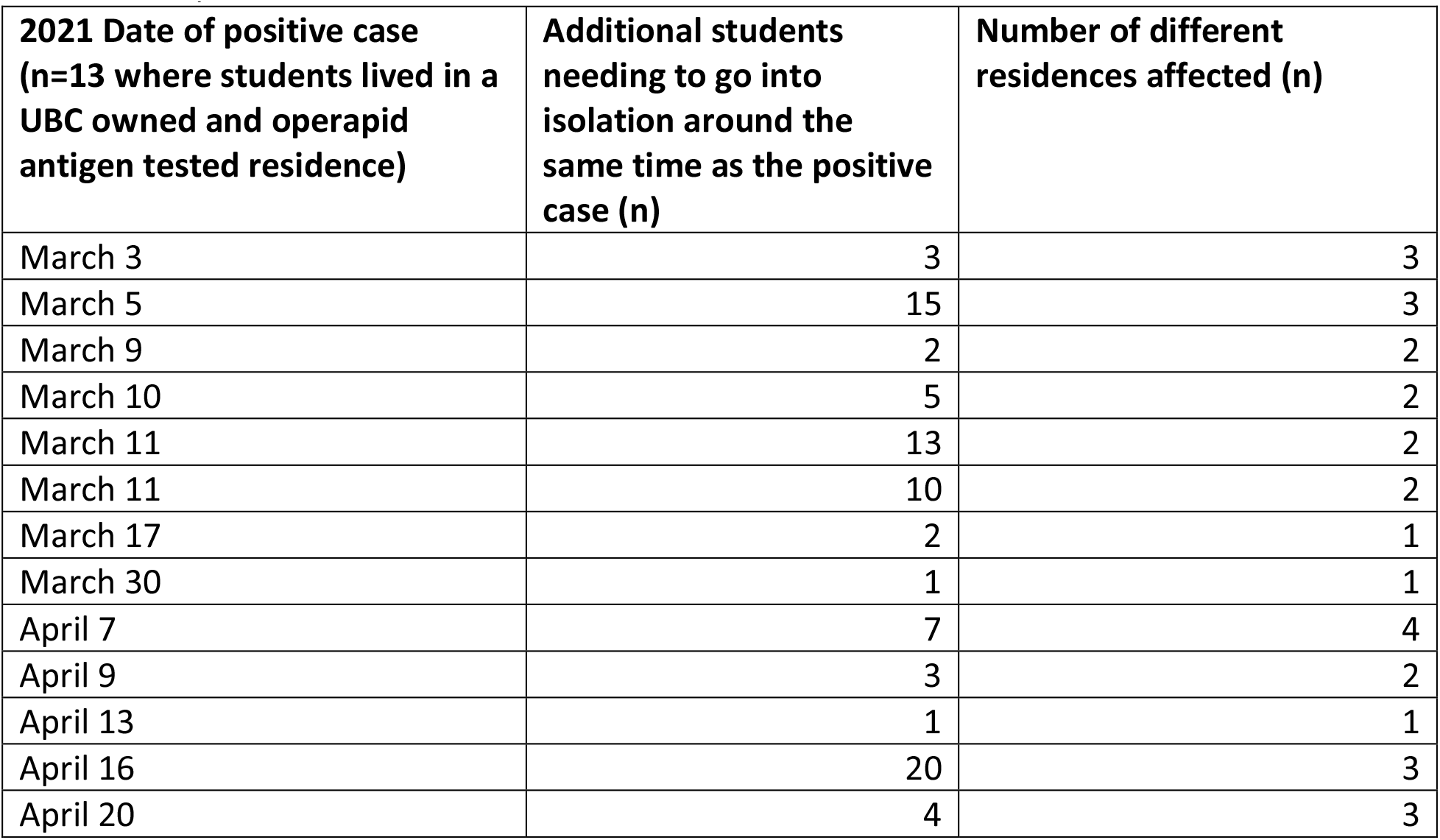
Additional students living in congregate housing who went into self-isolation around the time the positive case was found

| **2021 Date of positive case**<br>**(n=13 where students lived in a**<br>**UBC owned and operated**<br>**antigen tested residence)** | **Additional students**<br>**needing to go into**<br>**isolation around the**<br>**same time as the positive**<br>**case (n)** | **Number of different**<br>**residences affected (n)** |
| --- | --- | --- |
| March 3 | 3 | 3 |
| March 5 | 15 | 3 |
| March 9 | 2 | 2 |
| March 10 | 5 | 2 |
| March 11 | 13 | 2 |
| March 11 | 10 | 2 |
| March 17 | 2 | 1 |
| March 30 | 1 | 1 |
| April 7 | 7 | 4 |
| April 9 | 3 | 2 |
| April 13 | 1 | 1 |
| April 16 | 20 | 3 |
| April 20 | 4 | 3 |

The early identification of the positive cases led to the identification of 6 different clusters where 5-16 additional cases per cluster were found. The nature of the clusters included those living in congregate housing (UBC operated student residences or students living on campus, such as fraternity and sorority houses) and varsity athletes. One of these clusters included students playing musical instruments.

**Figure 1.**
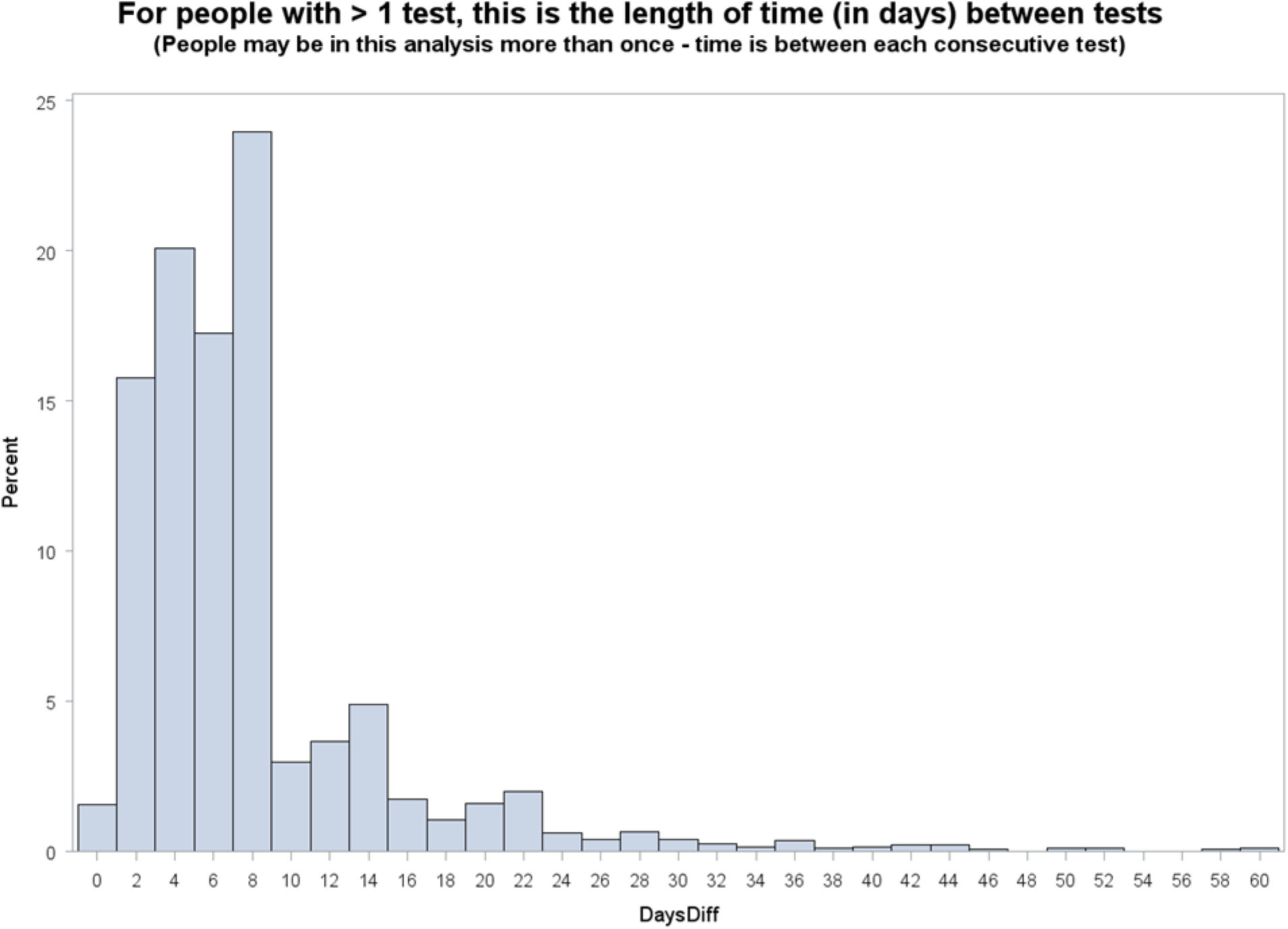

### Survey data

The survey ran for three weeks (March 24-31 and April 7-23, 2021). A total of 223 out of 601 screening site participants completed a survey. Table 3 shows the demographic characteristics of participants, who were mainly female and identified as women. Survey participants reported having a diversity of ethnic origins, including Chinese, Japanese, Korean, East Indian, Filipino, Vietnamese, Kenyan, Ugandan, Indigenous, and Latin American. Most survey participants were of European descent (45%) followed by East Asian (18%). Most participants lived at UBC (96%) in one of the UBC student residences (82%).

**Table 3.** Demographic characteristics of Orchard Commons survey participants.

| **Demographic characteristics**<br>**(n=223)** | **%** |
| --- | --- |
| **Sex** |  |
| Female | 60 |
| **Gender\*** |  |
| Women | 59 |
| Men | 37 |
| **Ethnicity** |  |
| Indigenous, Metis, Inuit | 3 |
| African/Caribbean | 5 |
| East Asian | 18 |
| South, Southeast and West Asian | 11 |
| European/Asian | 9 |
| South American | 3 |
| European | 45 |
| Other | 2 |
| **Housing** |  |
| At UBC | 96 |
| Off campus | 6 |
| UBC owned and operated student residence | 82 |
| Privately owned and operated residence on campus | 12 |

\*2% self identified as non-binary and other

Half of all survey participants reported they had low risk, very low risk, or no chance of getting COVID-19. Forty percent reported having some chance of getting COVID-19; 1 in 10 reported having a high chance of getting COVID-19. All survey participants stated they thought their rapid test result would be negative. Reasons provided as to why they thought their results would be negative were for the most part related to carefully following provincial health guidelines and protocols such as washing their hands, wearing a mask, avoiding crowds, and staying within their small social “bubble”. The vast majority of participants also stated that they had no to minimal interactions or recent exposure with someone who tested positive. Having no symptoms and having tested negative in the past were also mentioned.

The overwhelming majority (98%) of survey participants found the rapid antigen testing acceptable/very acceptable and 97% reported they would come back again for another test. In addition to wanting to play their part in research and data collection, the majority said they would return for testing because they wanted to be safe, confirm the belief that they were negative, to avoid the spread of COVID-19 (especially mentioned by varsity athletes), and to protect others. Many mentioned that it provided them with peace of mind and reduced their stress/anxiety level. More than two thirds of the respondents highlighted easy access to the rapid testing (e.g., obviated the need to take public transport to a dedicated community-based collection site), that it was quick, simple, painless, and free.

Some participants mentioned the low prevalence of positive cases and the perceived diminished accuracy of the rapid testing, stating for example “I have heard somewhere that basically no tests were coming back positive, though - there’s some worry that the test doesn’t actually catch Covid most times.” Finally, some staff mentioned wanting to protect themselves, others (family), and “… I want to be an example of best practice for anyone working here at UBC.”

## Discussion

We successfully deployed rapid antigen testing using the BD Veritor COVID-19 RAT in an asymptomatic sample of university students and staff living or working in congregate housing. Importantly, 55% of participants engaged in more frequent testing. Regular and repeated testing using rapid antigen tests can increase sensitivity in detecting active COVID-19 cases. This kind of COVID-19 screening program can potentially interrupt transmission chains and reduce overall community spread.(16) While the use of the screening site was voluntary, themes align with what Crozier, et al (21) explain as “testing to protect.” Survey participants stated they would return because they wanted to protect themselves and others. Providing accessibility of easy, pain-free testing to people living in congregate housing – as well as supports to self isolate – are important strategies used by the university to detect and isolate COVID-19 in the early stages of infection.

This work is a first in a Canadian university setting and targeting a sample of asymptomatic people living in congregate housing. The survey results are consistent with past work that females more often seek out healthcare.(22) While the vast majority of those who tested were negative, the majority of COVID-19 positives diagnosed through our site were males. Increased access to rapid testing is especially important for otherwise healthy young males who may not necessarily seek out health care as often as their young female counterparts.

Our results suggest there was an absence of harm related to false positives. While there was inconvenience of self-isolation, the confirmatory test was provided to participants within 7-10 hours of testing. Students may have missed some of their classes but the procedures associated with a presumptive (rapid antigen test) positive did not impede their schoolwork. Indeed, we found for some, serial testing provided psychological benefit by providing peace of mind.

Clinical, or diagnostic tests, such as the PCR are designed for people with symptoms and require high analytic sensitivity.(16) Rapid antigen tests such as the BD Veritor are fundamentally different in that they can be used for effectively detecting COVID-19 during the period of infectivity. The utility of these tests is to rapidly identify infectious cases of COVID-19 (including asymptomatics), and then to have these individuals placed in isolation, thereby interrupting chains of transmission and assisting in reducing community prevalence of what can be a debilitating infection. As countries continue to loosen COVID-19 public health restrictions, and as more people receive vaccinations, rapid antigen testing can be an additional layer of protection for individuals and for the broader community. These tests can be used in workplaces or events with inherent risk for contracting COVID-19 (e.g. airports, conferences, cultural events, food processing plants). They could also be offered to those living in congregate housing where there are shared amenities or sleeping quarters (e.g. university/college residences, migrant farm workers’ housing, long term care).

This work is limited in that this was a convenience sample of those who lived or worked at a university and chose to come to the screening site. The response rate to the survey was low. The strength of this work is the implementation and increased access of a rapid antigen testing site for those who were asymptomatic and living in congregate housing where there are no mandatory testing rules.

Rapid antigen testing as a screening intervention to detect COVID-19 is feasible to implement amongst people who are asymptomatic and live or work in a university residence. The ability to break silent chains of transmission early is a positive population health benefit since these people and any structural supports provided to them (e.g. self-isolation accommodation) could then prevent transmission to others. Rapid testing is relatively inexpensive (<$5), returns results quickly to limit asymptomatic spread and easy to execute to allow for frequent testing. More widespread implementation of rapid testing may help mitigate personal, healthcare and economic damage caused by COVID-19.

## Research in context panel

### Evidence before this study

Testing remains a cornerstone of mitigation and containment strategies, with recent data showing that one third of people infected with COVID-19 may be asymptomatic. A 2021 systematic review completed by Dinnes and colleagues suggests that rapid antigen tests can be used to detect COVID-19 amongst individuals who are infectious. These lateral flow assays provide increased access to results at the point of care at a lower cost and greatly improve accessibility and turnaround time making them appealing to industry, policy makers, and healthcare professionals. Yet, there is currently no universal public health policy for systematic COVID-19 screening for those who are asymptomatic or within specific contexts such as living in congregate housing.

### Added value of this study

Rapid antigen testing was successfully deployed in an asymptomatic sample of university students and staff living or working in congregate housing. The majority of participants engaged in serial testing which can increase sensitivity in detecting infectious COVID-19 cases. This kind of COVID-19 screening program can potentially interrupt silent transmission chains and reduce overall community spread. This work is a first in a Canadian university setting and targeting a sample of asymptomatic people living in congregate housing.

### Implications of all the available evidence

Access to rapid antigen testing provides an important strategy that can be used by the post-secondary institutions and other places that provide congregate housing to detect and isolate COVID-19 in the early stages of infection.

## Data Availability

De-identified data are available upon request, after peer reviewed publication.

## Acknowledgements

UBC rapid testing advisory group. We would like to acknowledge the students and staff at UBC who participated, UBC School of Nursing faculty and staff who donated their time to administer the testing. This work was made possible by the donation of rapid antigen tests by the Safe Restart Agreement Contribution Program Secretariat (testing, contact tracing and data management).

## Author contributions

Conceptualization, data curation, methodology, analysis, resources: Wong, S.T., Romney, M.G., Matic, N., Saewyc, E., Schwandt, M., Sin, D.D.

Data curation, supervision, writing, project administration: Haase, K., Ranger, M., Dhari, R., Affleck, F., Tan, E., Ndateba, I., Tobias, E.,

All authors provided input into the original draft and writing, review and editing.

## Access to the study data

all authors had full access to all the data in the study and accept responsibility to submit for publication

